# ChineseCVD: first-in-world, web-based, Chinese-specific Cardiovascular Risk Calculator incorporating long COVID, COVID-19 vaccination, SGLT2i and PCSK9i treatment effects

**DOI:** 10.1101/2023.10.27.23297656

**Authors:** Carlin Chang, Gary Tse, Quinncy Lee, Oscar Hou In Chou, Teddy Tai Loy Lee, Bosco Kwok-hei Leung, Amy Lee Ngai, Adan Khan, Wing Tak Wong, Abraham Ka Chung Wai, Kang-yin Chen, Tong Liu, Jiandong Zhou

## Abstract

**Background:** Web-based risk prediction tools for cardiovascular diseases are crucial for providing rapid risk estimates for busy clinicians, but there is none available specifically for Chinese subjects. This study developed ChineseCVD, first-in-world web-based Chinese-specific Cardiovascular Risk Calculator incorporating long COVID, COVID-19 vaccination, SGLT2i and PCSK9i treatment effects.

**Methods:** Adult patients attending government-funded family medicine clinics in Hong Kong between 1^st^ January 2000 and 31^st^ December 2003 were included. The primary outcome was major adverse cardiovascular events (MACE) defined as a composite of myocardial infarction, heart failure, transient ischaemic attacks/ischaemic strokes, and cardiovascular mortality.

**Results:** A total of 155,066 patients were used as the derivation cohort. Over a median follow-up of 16.1 (11.6-17.8) years, 31,061 (20.44%) had MACE. Cox regression identified male gender, age, comorbidities, cardiovascular medications, systolic and diastolic blood pressure, and laboratory test results (neutrophil-lymphocyte ratio, creatinine, ALP, AST, ALT, HbA1c, fasting glucose, triglyceride, LDL and HDL) as significant predictors of the above outcomes. ChineseCVD further incorporates the impact of smoking status, COVID-19 infection, number of COVID-19 vaccination doses, and modifier effects of newest medication classes of PCSK9i and SGLT2i. The calculator enables clinicians to demonstrate to patients how risks vary with different medications.

**Conclusions:** The ChineseCVD risk calculator enables rapid web-based risk assessment for adverse cardiovascular outcomes, thereby facilitating clinical decision-making at the bedside or in the clinic.

## Introduction

Cardiovascular diseases (CVDs), including myocardial infarction, heart failure, transient ischaemic attacks/ischaemic strokes, are leading causes of mortality, morbidity, and lower quality of life globally. Risk prediction tools or calculators are crucial for providing rapid risk estimates for busy clinicians, guiding decision making and clinical management at both primary and secondary care levels. The most popular CVD calculators include ASCVD Risk Estimator + created by the American College of Cardiology, Framingham Risk Score and QRisk3, but others also exist, including Reynolds Risk Score ^1^, Pan American Health Organization Cardiovascular Risk Calculator ^2^, AusCVDRisk ^3^, New Zealand cardiovascular risk prediction equations ^4^.

However, as noted in an Editorial by our team published in 2018 ^5^, there is no tool that is tailored and individualised specifically for Chinese subjects. Since 2020, we have been conducting population-based studies evaluating risk factors for CVDs, with a focus on blood pressure ^6,7^, lipid ^8^ and glycemic tests ^9^, as well as measures of their visit-to-visit variability, for risk prediction for myocardial infarction, stroke, heart failure, dementia ^10^ and other outcomes such as anxiety ^11^ and bone fractures ^12^. The ultimate goal has been the creation and development of risk models that are enhanced by AI. Our unique approach is personalised care, with individualised risk prediction based on different diseases. The first-in-world Chinese-specific CVD risk models were published by other teams ^13-16^. To further enhance risk prediction using state-of-the-art AI technology, our team recently reported on the development of PowerAI-CVD, which is the first Chinese-specific, validated artificial intelligence-powered *in-silico* predictive model for CVD ^17^. The model was developed from large historical cohort of Hong Kong Chinese patients attending publicly managed family medicine clinics between 2000 and 2003 with two decades of follow-up data available for myocardial infarction, heart failure and transient ischaemic attacks/ischaemic strokes separately, as well as their composite outcome (major adverse cardiovascular events), at 1-, 3-, 5-, 10- and 20-year time points.

Whilst the enhancement by AI can significantly improve the predictive performance and it can be embedded in health record systems in the form of a dashboard, the complexity of the algorithm represents a barrier for implementation in routine clinical settings. By contrast, a web-based calculator has the capability of being easily accessible and can be used by busy clinicians by the bedside or in the clinic. In 2020, our team was amongst the first to develop a Chinese-specific risk calculator for prediction of severe COVID-19 disease, which was made freely available on QxMD ^18^. In this study, we present a Chinese-specific risk calculator based on these published findings. The major novelty of our model is the incorporation of factors include prior COVID-19 infection, COVID-19 vaccination and consideration of treatment effects including the latest drug class such as proprotein convertase subtilisin/kexin type 9 (PCSK9) inhibitors.

## Methods

### Ethics approval and study cohort

This study was approved by the Institutional Review Board of the University of Hong Kong/Hospital Authority Hong Kong West Cluster Institutional Review Board (HKU/HA HKWC IRB) (UW-20-250 and UW 23-339) and The Joint Chinese University of Hong Kong (CUHK) Hospital Authority New Territories East Cluster (NTEC) Clinical Research Ethics Committee (CREC) (2018.309 and 2018.643). It also complied with the Declaration of Helsinki.

This was a retrospective population-based study of prospectively collected electronic health records using the Clinical Data Analysis and Reporting System (CDARS) managed by the Hong Kong Hospital Authority (HA). These records include information from public hospitals, their affiliated outpatient clinics, day-care centres and ambulatory care facilities. This system has been used extensively by research groups from Hong Kong ^18-20^.

The procedures for data extraction and data analysis has been described in the previous study ^17^. Briefly, the inclusion criteria were patients attending family clinics managed by the Hospital Authority of Hong Kong between 1^st^ January 2000 and 31^st^ December 2003. The exclusion criteria were patients who died within 30 days of the baseline date or who were <18 years old. The following clinical information was extracted during the baseline period, defined as 1^st^ January 2000 to 31^st^ December 2003: demographic details (age and gender), blood pressure (mean systolic blood pressure [SBP] and diastolic blood pressure [DBP]), existing diseases of diabetes mellitus, hypertension, chronic obstructive pulmonary disease, ischaemic heart disease, heart failure, myocardial infarction, and stroke/TIA, prescription details of different cardiovascular drugs. The diseases were defined using International Classification of Diseases (ICD)-9 codes (**Supplementary Appendix: Table 1**). Complete blood count, liver and renal function tests, glycemic and lipid tests during the baseline period were extracted.

### Outcomes and statistical analysis

The primary outcome was MACE, defined as any of the following events: myocardial infarction, heart failure, TIA/stroke and cardiovascular mortality, with follow-up until 31^st^ December 2019. Continuous variables were presented as median (95% confidence interval [CI] or interquartile range [IQR]) and categorical variables are shown as frequency (%). The Mann-Whitney U test was used to compare continuous variables. The χ2 test with Yates’ correction was used for 2×2 contingency data, and Pearson’s χ2 test was used for contingency data for variables with more than two categories. All statistical tests were two-tailed and considered significant with P <0.05. Analyses were conducted using RStudio software (Version: 1.1.456) or Python (Version: 3.6).

### Development of predictive models and validation

To develop the ChineseCVD model, we followed the approach taken by the pooled cohort equation model, where we used a historical cohort for derivation followed by a contemporary cohort for validation ^21^. Cox regression was used to identify significant predictors of MACE. The natural logarithm values of the hazard ratios were used for model development, with 3-, 5-, 10- and 20-year risks estimated using cohort-level event rates at these time points. For validation of 5-year CVD risk, an independent cohort of patients attending family medicine clinics managed by the Hong Kong West Cluster (HKWC) of the Hospital Authority in 2019, excluding patients in the 2000 to 2003 cohort used for model development, will be used. The model will then be recalibrated and tested using another independent cohort of patients attending family medicine clinics managed by clusters other than HKWC.

## Results

### Model development and validation

In the development cohort, a total of 155066 patients were included. Of these, 31,061 (20.44%) patients had MACE over a median follow-up of 16.1 (11.6-17.8) years. Cox regression identified age, male gender, SBP, DBP, diabetes mellitus, hypertension, hypertension treatment with antihypertensive, COPD, IHD, IHD treatment with antiplatelet, MI, HF, AF, AF treatment with anticoagulant, TIA/stroke, NLR, creatinine, ALP, ALT, triglyceride, LDL, HDL, HbA1c and fasting glucose as significant predictors. The risks of smoking status were obtained from the analysis of the China Kadoorie Biobank ^22^. To incorporate the novel variables of prior COVID-19 infection ^23^, number of COVID-19 vaccination doses ^24^ and initiation of statins ^25^, SGLT2i ^26^ or PCSK9i ^27^ therapy, HRs were identified from these published studies and added to our model, termed ChineseCVD. The corresponding beta values are shown in **Table 1** and a graphical interface of ChineseCVD is shown in **Figure 1**. Validation for 5-year CVD risk using an independent cohort of patients attending family medicine clinics managed by the Hong Kong West Cluster of the Hospital Authority in 2019 will be performed. The model will then be recalibrated and tested using another independent cohort of patients attending family medicine clinics managed by clusters other than HKWC.

**Table 1.** Univariable Cox regression to predict MACE in patients attending family medicine clinics.

| Characteristics | MACE | Beta coefficient |
| --- | --- | --- |
|  | HR [95% CI];P value |  |
| **Demographics** |  |  |
| Male gender | 1.13 [1.11-1.16] | 0.122217633 |
| Baseline age, year | 1.04 [1.04-1.05] | 0.039220713 |
| BP (Systolic)-mean | 1.029 [1.029-1.030] | 0.028587457 |
| BP (Diastolic)-mean | 0.979 [0.978-0.981] | -0.021223636 |
| Current smoker (male) | 1.28 [1.23-1.34] | 0.246860078 |
| Ex-smoker (male) | 1.06 [0.99-1.14] | 0.058268908 |
| Current smoker (female) | 1.17 [1.10-1.26] | 0.157003749 |
| Ex-smoker (female) | 1.09 [0.93-1.26] | 0.086177696 |
| **Comorbidities** |  |  |
| Diabetes mellitus | 2.75 [1.99-3.80] | 1.011600912 |
| Hypertension | 3.47 [3.35-3.60] | 1.244154594 |
| Chronic obstructive pulmonary disease | 4.67 [4.12-5.30] | 1.541159072 |
| Ischemic heart disease | 4.04 [3.81-4.29] | 1.396244692 |
| Heart failure | 15.44 [14.42-16.54] | 2.736961545 |
| Myocardial infarction | 4.39 [3.70-5.21] | 1.479329227 |
| Atrial fibrillation | 9.25 [8.56-10.00] | 2.224623552 |
| Transient ischaemic attack/Stroke | 4.64 [4.23-5.09] | 1.534714366 |
| Antihypertensive treatment | 0.66 [0.63-0.68] | -0.419979151 |
| Antiplatelet treatment | 0.82 [0.78-0.87] | -0.193272388 |
| Anticoagulant treatment | 0.25 [0.22-0.25] | 2.224623552 |
| SGLT2i treatment | 0.86 [0.80-0.93] | -0.15082289 |
| PCSK9i treatment | 0.59 [0.37-0.92] | -0.527632742 |
| Any COVID-19 disease | 1.12 [1.05-1.19] | 0.113328685 |
| Severe COVID-19 disease | 1.99 [1.04-3.76] | 0.6886325 |
| COVID-19 vaccination (1 dose) | 0.79 [0.71-0.88] | -0.235722334 |
| COVID-19 vaccination (2 doses) | 0.63 [0.58-0.69] | -0.46203546 |
| COVID-19 vaccination (3 doses) | 0.59 [0.54-0.66] | -0.527632742 |
| **Laboratory tests** |  |  |
| Neutrophil-to-lymphocyte ratio | 1.022 [1.020-1.025] | 0.021761492 |
| Creatinine, umol/L | 1.003 [1.003-1.003] | 0.002995509 |
| Alkaline phosphatase, U/L | 1.001 [1.001-1.001] | 0.0009995 |
| Alanine transaminase, U/L | 0.999 [0.998-0.999] | -0.0010005 |
| Mean triglyceride, mmol/L | 0.97 [0.95-0.98] | 1.8494 |
| Mean low-density lipoprotein, mmol/L | 0.69 [0.67-0.70] | 2.5412 |
| Mean high-density lipoprotein, mmol/L | 0.53 [0.51-0.55] | 1.1338 |
| Mean HbA1C, % | 1.08 [1.06-1.10] | 7.9857 |
| Mean fasting glucose, mmol/L | 1.08 [1.07-1.08] | 5.6113 |

**Figure 1.**
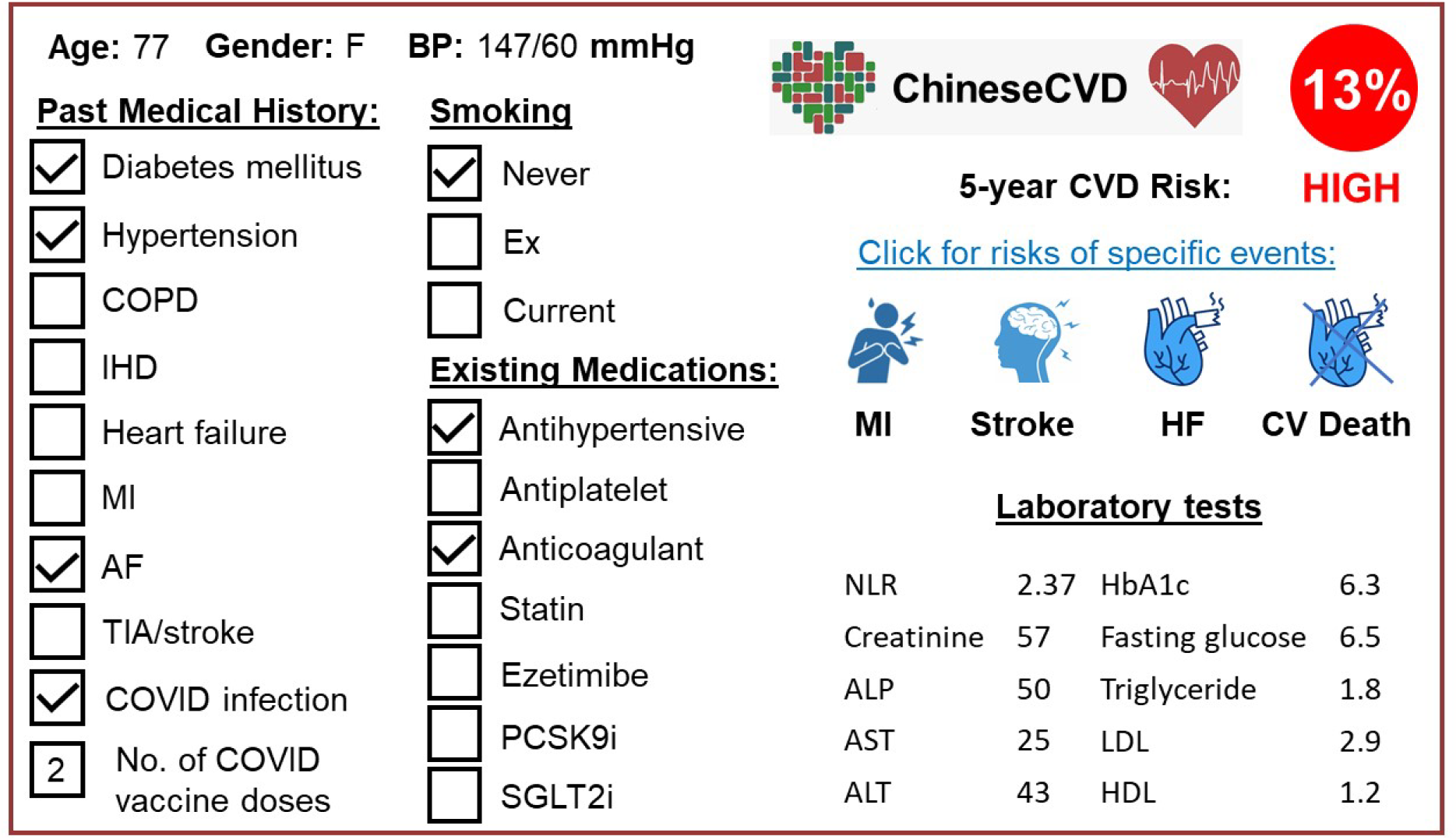
ChineseCVD Web calculator for instantons calculation of 5-year MACE risk.

## Discussion

The main findings of this study are the development of ChineseCVD, a web-based risk prediction tools freely available to clinicians and investigators worldwide for predicting incident cardiovascular events. The novelty is that our model incorporates new variables of previous COVID-19 infection, the number of COVID-19 vaccination doses, and treatment effects of cardiovascular medications, such as statins and PCSK9i, and the newest antidiabetic medication class, SGLT2i.

A number of risk calculators are currently available for cardiovascular risk prediction, including QRISK3® ^28^, developed from the QResearch database that contains data from General practices in England, and ACC/AHA Pooled Cohort Cardiovascular Risk Equations, developed from primary data of studies published in the pre-statin era, including the Atherosclerosis Risk In Communities study ^29^, Cardiovascular Health Study ^30^, Coronary Artery Risk Development in Young Adults study ^31^, which were combined with data from the Framingham ^32^ and Framingham Offspring ^33^ studies. By contrast, there are few risk scores developed specifically for Asian populations and yet Asians different from Western individuals in terms of genetics, culture, lifestyle, social and behavioural characteristics ^34^. Relevant for cardiovascular risk, the Asia Pacific Cohort Studies Collaboration found higher systolic blood pressure, total cholesterol, and cardiovascular event rates in the Framingham cohort compared to six cohort studies from Japan, Korea and Singapore, and six cohort studies from China ^35^. Given these differences, direct application of Western risk models will inevitably lead to inaccuracies in the risk estimates ^36^. Therefore, our team recently developed the first Chinese-specific, validated artificial intelligence-powered *in-silico* predictive model for cardiovascular disease, called PowerAI-CVD ^17^. Comparing different artificial intelligence approaches, we found that CatBoost significantly outperformed XGBoost, Gradient Boosting, Multilayer Perceptron, Random Forest, Naive Bayes, Decision Tree, k-Nearest Neighbor, AdaBoost, SVM-Sigmod and logistic regression, showing a classification accuracy of. Strikingly, it showed significant prediction performance across different subgroups of age, sex and both for primary and secondary prevention purposes.

To facilitate implementation, this study proposes a simplified, score-based risk calculator, ChineseCVD, based on these validated risks. The novelty of ChineseCVD is the consideration of treatment effects, specifically statins and new drug classes of PCSK9i and SGLT2i. Our model also incorporated the impact of long COVID on the cardiovascular system, as well as the protective effects of COVID-19 vaccination on MACE. These features deal with the limitations of existing risk calculators, where the scores do not consider initiation of antiplatelet drugs, statins and antihypertensive medications. Current risk models have the limitation of overestimating CVD risk, which is partly attributed to the lack of statin use in historical cohort years from which the models were developed.

ChineseCVD is specifically designed for the primary care settings, as the model development had utilised family medicine clinic data. Our unique approach is personalised care, with individualised risk prediction based on different diseases and outcomes. Thus, we have developed CVD risk models for diabetes mellitus ^37-39^, which captured data from both primary and secondary care settings. The first version, PowerAI-Diabetes, third-in-world, Chinese-specific AI-driven predictive model for predicting diabetic complications and first-in-world to incorporate lipid and glycaemic variability with AI, was recently developed ^40^. We are incorporating new information such as treatment effects of first- and second-line anti-diabetic medications, such as metformin, sulphonylureas ^41,42^, SGLT2 inhibitors, DPP4 inhibitors, GLP1 agonists ^43,44^, which would impact on the risks of CVD and other adverse events. Our AI-enhanced diabetes model complement previously published, Chinese-specific diabetes CVD risk models by other groups ^45,46^. Our future model will also capture fluctuations in inflammation using biomarkers ^47^. Finally, we are developing cancer-specific CVD risk models based on our recent publications ^48^, most notably our series on prostate cancer ^49-53^.

## Conclusions

The ChineseCVD risk calculator enables rapid web-based risk assessment for adverse cardiovascular outcomes, thereby facilitating clinical decision-making at the bedside or in the clinic.

## Data Availability

All data produced in the present study are available upon reasonable request to the authors

